# Investigating the Role of Neighborhood Socioeconomic Status and Germline Genetics on Prostate Cancer Risk

**DOI:** 10.1101/2024.07.31.24311312

**Authors:** Jonathan Judd, Jeffrey P. Spence, Jonathan K. Pritchard, Linda Kachuri, John S. Witte

## Abstract

**Background:** Genetic factors play an important role in prostate cancer (PCa) development with polygenic risk scores (PRS) predicting disease risk across genetic ancestries. However, there are few convincing modifiable factors for PCa and little is known about their potential interaction with genetic risk. We analyzed incident PCa cases (n=6,155) and controls (n=98,257) of European and African ancestry from the UK Biobank (UKB) cohort to evaluate the role of neighborhood socioeconomic status (nSES)–and how it may interact with PRS–on PCa risk.

**Methods:** We evaluated a multi-ancestry PCa PRS containing 269 genetic variants to understand the association of germline genetics with PCa in UKB. Using the English Indices of Deprivation, a set of validated metrics that quantify lack of resources within geographical areas, we performed logistic regression to investigate the main effects and interactions between nSES deprivation, PCa PRS, and PCa.

**Results:** The PCa PRS was strongly associated with PCa (OR=2.04; 95%CI=2.00-2.09; P<0.001). Additionally, nSES deprivation indices were inversely associated with PCa: employment (OR=0.91; 95%CI=0.86-0.96; P<0.001), education (OR=0.94; 95%CI=0.83-0.98; P<0.001), health (OR=0.91; 95%CI=0.86-0.96; P<0.001), and income (OR=0.91; 95%CI=0.86-0.96; P<0.001). The PRS effects showed little heterogeneity across nSES deprivation indices, except for the Townsend Index (P=0.03).

**Conclusions:** We reaffirmed genetics as a risk factor for PCa and identified nSES deprivation domains that influence PCa detection and are potentially correlated with environmental exposures that are a risk factor for PCa. These findings also suggest that nSES and genetic risk factors for PCa act independently.

## INTRODUCTION

Prostate cancer (PCa) is the second most common cancer in men in the United States, with the highest incidence rates observed in African American men (1). In addition to older age, there is evidence for a strong genetic component in PCa etiology, including family history and germline susceptibility loci (2). Genome-wide association studies (GWAS) have identified 451 independent risk variants for PCa, and these findings have been used to develop a polygenic risk score (PRS). Although the most recent PRS for PCa has improved risk prediction across ancestry groups, performance in African and East Asian populations continues to lag behind European ancestry (3).

Individual and neighborhood socioeconomic status (SES) has been explored as a potential risk factor for PCa (4–6), however, studies of neighborhood SES (nSES) have produced mixed results (7–9). Some find a positive relationship between nSES and PCa risk, theorizing that advantaged populations are more likely to undergo PSA screening or engage with the healthcare system (9). Other research has found the opposite effect with lower nSES groups being at higher risk, suggesting a target for public health interventions (8,10).

Gene-by-environment (GxE) interactions are thought to play an important role in complex trait variation across populations (11). In addition to interactions between environmental factors and specific risk variants, there is accumulating evidence that PRS performance is context-dependent and may be modulated by SES factors (12).

In this study, we investigated the main effects and interactions of PCa PRS and nSES using the UK Biobank (UKB), a recently published PCa PRS, and the English Indices of Deprivation.

## METHODS

### Study Population

The United Kingdom Biobank (UKB) is a prospective study of 503,317 participants aged 40–69 years across England, Wales and Scotland (13). Between 2006 and 2010, the UKB collected genotyping data, linkage to national cancer registries, and demographic and questionnaire data at 22 assessment centers.

All quality control and phenotyping of data used in our analyses was described and performed in previous studies (14). We restricted our analyses to men with concordant self-reported and genetically inferred sex, available genotyping data, known values for all deprivation indices, and known cancer status histories. Additionally, we excluded men with a cancer diagnosis before assessment were or a post-assessment cancer diagnosis other than PCa or non-melanoma skin cancer. Controls included men without any cancer diagnosis (excluding non-melanoma skin cancer). Analyses were performed separately in populations groups defined based on self-reported ethnicity and genetic principal component (PCs) values within five standard deviations of the UK10K and 1000 Genomes phase 3 reference panel group means. Using this criteria, we analyzed two separate populations throughout this study that we will refer to as the European (5,960 cases and 93,990 controls) and African (109 cases and 1,226 controls) ancestries (14,15).

### Indices of Deprivation & Townsend Index

The English Indices of Deprivation are a series of metrics calculated by the UK Government that assess relative deprivation of geographic areas across seven domains: Income Deprivation; Employment Deprivation; Health Deprivation and Disability; Education, Skills and Training Deprivation; Barriers to Housing and Services; Living Environment Deprivation; and Crime (16). Domain-specific indices are combined into a single Index of Multiple Deprivation (IMD) using predefined weights: Income=22.5%; Employment=22.5%; Health and Disability=13.5%; Education, Skills, and Training=13.5%; Housing and Services=9.3%; Environment=9.3%; Crime=9.3% (17).. The Townsend Index is another nSES deprivation metric composed of four area-based indicators: percentage of households without a car, overcrowded households, households not owner-occupied, and persons unemployed, that are standardized and summed (18). For all UKB participants, values of deprivation indices were assigned based on their postcode and year of assessment. For detailed descriptions, see Technical Report and Section 3.2 of (17,18), respectively.

### Prostate Cancer PRS

The PCa PRS used in this analysis consists of 269 risk variants identified using multi-ancestry fine-mapping and summary statistics from a GWAS of 107,247 cases and 127,006 controls across three genetic ancestries (19,20). UKB participants were not included in the GWAS meta-analysis used for training this PRS model. For each individual, 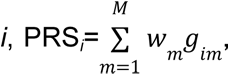 where *g*_im_ is the genotype dosage of variant *m* of *M* total variants for individual *i* and *w_m_* is a variant-specific weight (logOR scale).

Associations with PCa were estimated separately in each ancestral population using logistic regression with adjustment for age at assessment and the top 10 genetic ancestry principal components (PCs). We estimated odds ratios (OR) per standard deviation (SD) increase in PRS_269_, standardized within each ancestry group. We also modeled the PRS as a discrete variable with cut-points based on deciles with the 40-60% group as the reference category.

Our primary analysis uses PRS_269_, which was not trained on the largest PCa GWAS to date but avoids overlap with UKB. We also present sensitivity analyses using two of the most recent PCa PRS models for which UKB was part of the training data (3): one model with 451 variants, and another developed using PRS-CSx (21).

### Modeling Neighborhood Socioeconomic Status

We used logistic regression to assess the potential association between nSES deprivation, as measured by the English Indices of Deprivation and the Townsend Index, and PCa. Age at assessment was included as a covariate for all models. Additionally, to investigate if any associations between nSES deprivation and PCa are mediated by PSA screening and healthcare utilization (22), we used the “Ever had a PSA test” variable in UKB and calculated a proxy for healthcare utilization by summing the number of general practitioner visits and hospital inpatient episodes in the year prior to assessment (23).

To evaluate the relationship between nSES deprivation, PSA screening, and PCa, we performed logistic regression between nSES deprivation and UKB’s “Ever had a PSA test” variable before modeling the association between nSES deprivation and PCa while adjusting for PSA screening and age. We repeated this analysis for healthcare utilization by modeling the association between nSES deprivation and number of clinical visits before analyzing the association between nSES deprivation and PCa while adjusting for number of clinical visits and age. Lastly, we calculated the correlation between “Ever had a PSA test”, number of clinical visits, and all nSES variables before estimating the association between nSES deprivation and PCa using PSA screening, number of clinical visits, and age as covariates.

Finally, to further investigate the potential impact of high nSES on increasing PSA screening and prostate cancer, we performed a stratified analysis based on self-reported screening status. We then performed a difference of means z-test for each nSES variable between the ever had a PSA test and never had a PSA test group. Due to small sample sizes in the other ancestry groups, we performed this analysis on only individuals of European ancestry.

### Evaluation of Genetic Effect Modification

We investigated whether the PRS_269_ associations with PCa were modified by nSES deprivation in two ways. First, we fitted logistic regression models that included the main effects and an interaction between PRS_269_ and nSES to estimate their effects on PCa risk. We assessed modification by looking at whether the interaction term was associated with PCa. Second, we evaluated the standardized PRS_269_ effect within quartiles of each nSES index. We computed Cochran’s Q to assess heterogeneity in the effect size of the PRS across quartiles. This analysis was limited to men of European ancestry due to smaller sample sizes in the African ancestry (109 cases and 1,226 controls.

## RESULTS

### Participants Characteristics

This study included a total of 6,069 PCa cases and 95,216 cancer-free controls (**Table 1).** Most participants were of European ancestry (5,960 cases and 93,990 controls), compared to African (109 cases and 1,226 controls) ancestry. In these populations, 30% and 24% of participants reported having had at least one PSA test prior to recruitment. For both ancestries, the neighborhood deprivation scores, including the index of multiple deprivation (IMD) and Townsend Index, were lower in the cases compared to the controls. In contrast, the median number of clinical visits the year before assessment was higher for cases than controls in both European (Controls:6; Cases:7) and African (Controls:7; Cases:9) ancestries.

**Table 1:**
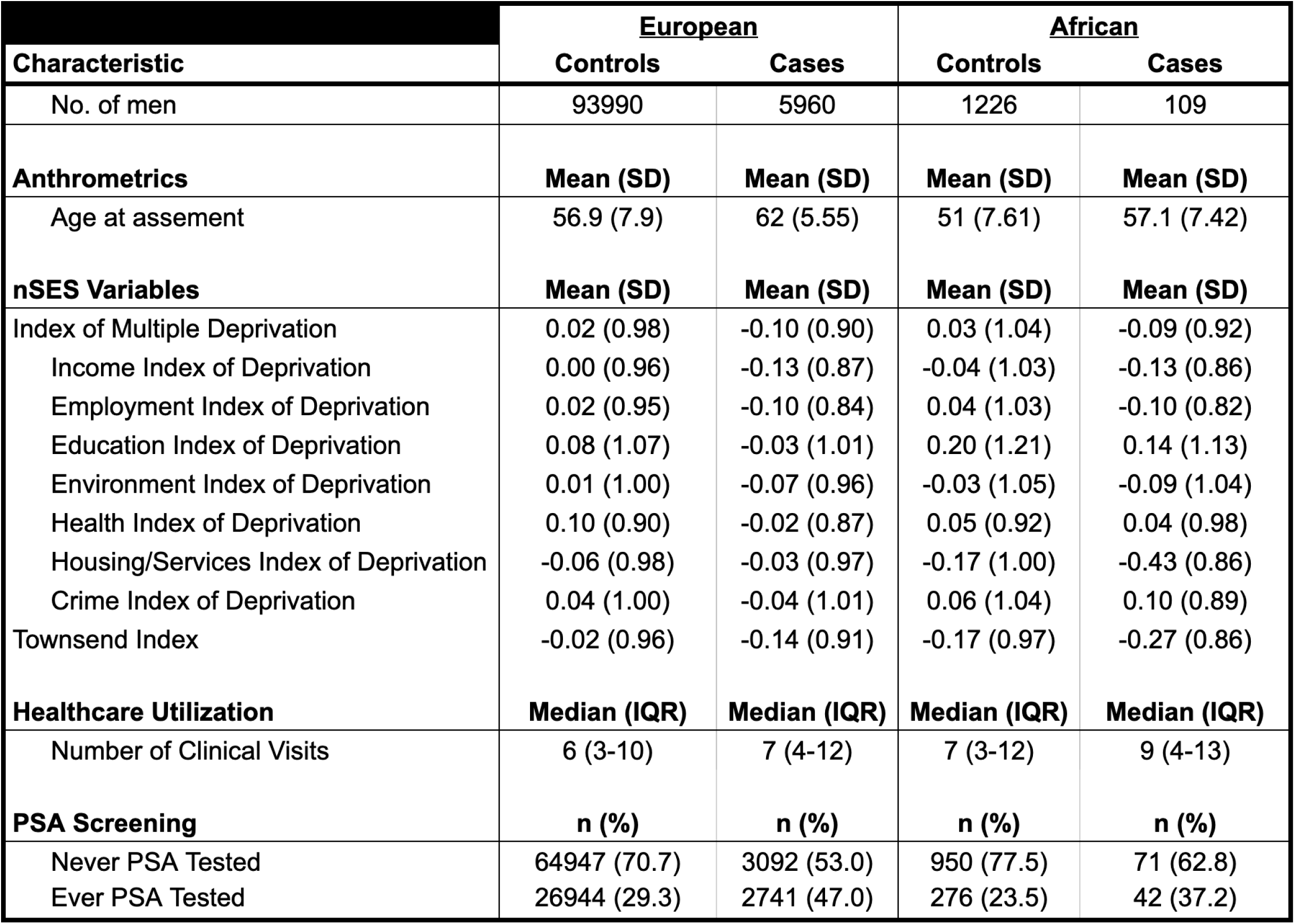
Baseline characteristics of men included in the analysis. Demographic and distribution characteristics for cases and controls for the European and African ancestries analyzed in the study. All values were obtained at time of assessment. Anthropometrics - Age, body mass index (BMI), and height (cm) - are non-standardized. Neighborhood socioeconomic status (nSES) deprivation variables include 2010 English Indices of Deprivation and Townsend Index. Index means and standard deviation (SD) are standardized for the total sample after stratifying on population, but before case/control status. Number of clinical visits is defined as the number of primary care and hospital inpatient visits per individual in the year prior to assessment. Median and interquartile range (IQR) for number of clinical visits is defined per population before stratifying on case/control status. PSA screening status per individual is determined by indicator for self-reported PSA screening at baseline (ever/never). Total number and percentages were calculated within both population and case/control status.

| **Characteristic** | **European** |  | **African** |  |
| --- | --- | --- | --- | --- |
|  | **Controls** | **Cases** | **Controls** | **Cases** |
| No. of men | 93990 | 5960 | 1226 | 109 |
| **Anthrometrics** | **Mean (SD)** | **Mean (SD)** | **Mean (SD)** | **Mean (SD)** |
| Age at assement | 56.9 (7.9) | 62 (5.55) | 51 (7.61) | 57.1 (7.42) |
| **nSES Variables** | **Mean (SD)** | **Mean (SD)** | **Mean (SD)** | **Mean (SD)** |
| Index of Multiple Deprivation | 0.02 (0.98) | -0.10 (0.90) | 0.03 (1.04) | -0.09 (0.92) |
| Income Index of Deprivation | 0.00 (0.96) | -0.13 (0.87) | -0.04 (1.03) | -0.13 (0.86) |
| Employment Index of Deprivation | 0.02 (0.95) | -0.10 (0.84) | 0.04 (1.03) | -0.10 (0.82) |
| Education Index of Deprivation | 0.08 (1.07) | -0.03 (1.01) | 0.20 (1.21) | 0.14 (1.13) |
| Environment Index of Deprivation | 0.01 (1.00) | -0.07 (0.96) | -0.03 (1.05) | -0.09 (1.04) |
| Health Index of Deprivation | 0.10 (0.90) | -0.02 (0.87) | 0.05 (0.92) | 0.04 (0.98) |
| Housing/Services Index of Deprivation | -0.06 (0.98) | -0.03 (0.97) | -0.17 (1.00) | -0.43 (0.86) |
| Crime Index of Deprivation | 0.04 (1.00) | -0.04 (1.01) | 0.06 (1.04) | 0.10 (0.89) |
| Townsend Index | -0.02 (0.96) | -0.14 (0.91) | -0.17 (0.97) | -0.27 (0.86) |
| **Healthcare Utilization** | **Median (IQR)** | **Median (IQR)** | **Median (IQR)** | **Median (IQR)** |
| Number of Clinical Visits | 6 (3-10) | 7 (4-12) | 7 (3-12) | 9 (4-13) |
| **PSA Screening** | **n (%)** | **n (%)** | **n (%)** | **n (%)** |
| Never PSA Tested | 64947 (70.7) | 3092 (53.0) | 950 (77.5) | 71 (62.8) |
| Ever PSA Tested | 26944 (29.3) | 2741 (47.0) | 276 (23.5) | 42 (37.2) |

### Relationship between PRS and Prostate Cancer

We reaffirmed the association between PRS_269_ and PCa using our specific sample (19). We found PRS_269_ to be associated with PCa in both the European (OR=2.04; 95%CI=2.00-2.09; P<0.001) and African (OR=1.35; 95%CI=1.16-1.58; P<0.001) ancestries. We also evaluated this association in two more recently developed PCa PRS models–one with 451 variants and the other using PRS-CSx, to investigate the change in the PRS and PCa association fueled by models from a larger more-diverse GWAS (3). When we did this, we found associations of similar magnitude across all three models for both populations (**Supp. Table 1**). Even though the secondary PRS models were developed from a larger GWAS than PRS_269_, they included UKB as part of their training data. Because this posed a risk of being overfit to our sample, PRS_269_ was used for all future analyses.

When we investigated if the relationship between PRS_269_ and PCa risk was linear, we found that the association increased across PRS deciles for men of European ancestry, but not men of African ancestry. Men in the top decile of the PRS distribution had a significantly increased risk of PCa compared to those with average PRS (40-60%) in both the European (OR=3.62; 95%CI=3.38-3.89; P<0.001) and African (OR=1.9;95%CI=1.13-3.20; P=0.02) ancestries. The association between PRS_269_ and PCa was statistically significant for all deciles in the European ancestry models, ranging from OR=0.26 to OR=3.62. However, in men of African ancestry, only the second highest decile of the PRS distribution had a significantly increased risk compared to the average (OR=1.76; 95%CI=1.04-3.00; P=0.04) **(Table 2)**. The associations in the African ancestry models were also less precise than those in the European models likely due to smaller samples, especially cases, in the lower decile groups.

**Table 2:** Odds ratios for prostate cancer by polygenic risk score (PRS) percentile stratified by genetic population. Odds ratios (OR) and 95%confidence intervals for the association between a specified Polygenic Risk Score 269 (PRS_269_) decile and Prostate Cancer (PCa) compared to individuals in the 40-60% percentile of scores (reference) for European and African ancestry samples. The number of cases and controls for each population and percentile group is given. PRS_269_ values were standardized per population before analysis.

| PRS Category | European |  |  | African |  |  |
| --- | --- | --- | --- | --- | --- | --- |
|  | Case/Control | OR (95% CI) | P-Value | Case/Control | OR (95 %CI) | P-Value |
| 0-10% | 203/15450 | 0.28 (0.25-0.33) | 4.52E-68 | 9/295 | 0.55 (0.26-1.18) | 1.24E-01 |
| 10-20% | 336/15254 | 0.51 (0.45-0.57) | 1.95E-31 | 13/280 | 0.91 (0.49-1.69) | 7.58E-01 |
| 20-30% | 453/15138 | 0.6 (0.54-0.67) | 1.77E-20 | 19/281 | 1.35 (0.76-2.38) | 3.07E-01 |
| 30-40% | 501/15040 | 0.73 (0.66-0.8) | 4.25E-10 | 22/263 | 1.05 (0.58-1.9) | 8.78E-01 |
| 40-60% | 1471/29540 | Reference | Reference | 34/549 | Reference | Reference |
| 60-70% | 953/14506 | 1.29 (1.19-1.41) | 3.58E-09 | 26/263 | 1.28 (0.72-2.26) | 4.06E-01 |
| 70-80% | 1148/14286 | 1.59 (1.46-1.72) | 6.27E-29 | 24/273 | 1.36 (0.79-2.36) | 2.73E-01 |
| 80-90% | 1425/13899 | 2.11 (1.95-2.28) | 2.76E-80 | 25/264 | 1.76 (1.04-3) | 3.64E-02 |
| 90-100% | 2299/12804 | 3.62 (3.38-3.89) | 1.04E-276 | 38/248 | 1.9 (1.13-3.2) | 1.49E-02 |

### Relationship between nSES and Prostate Cancer

For each ancestry, we assessed the relationship between nSES deprivation and PCa using a minimally adjusted model. In the European ancestry, six of the nine nSES deprivation indices had inverse associations with prostate cancer: Education (OR=0.92; 95%CI=0.90-0.95; P<0.001), Employment (OR=0.91; 95%CI=0.88-0.93; P<0.001), Health (OR=0.91; 95%CI=0.88-0.93; P<0.001), Income (OR=0.91; 95%CI=0.89-0.94; P<0.001), IMD (OR=0.92; 95%CI=0.89-0.95; P<0.001), and Townsend Index (OR=0.93; 95%CI=0.90-0.96; P<0.001) (all ORs for a SD unit increase in the deprivation index) (**Figure 1a, Supp. Table 2**). These results signify that higher deprivation and lower neighborhood resources are associated with lower PCa risk. In the European ancestry, the Housing/Services index (OR=1.04; 95%CI=1.01-1.06; P<0.001) was the only nSES domain with a positive association, and we found no associations between the Crime and Environment indices in this group. In men of African ancestry, none of the nSES deprivation indices were associated with PCa (**Figure 1b, Supp. Table 2**).

**Figure 1:**
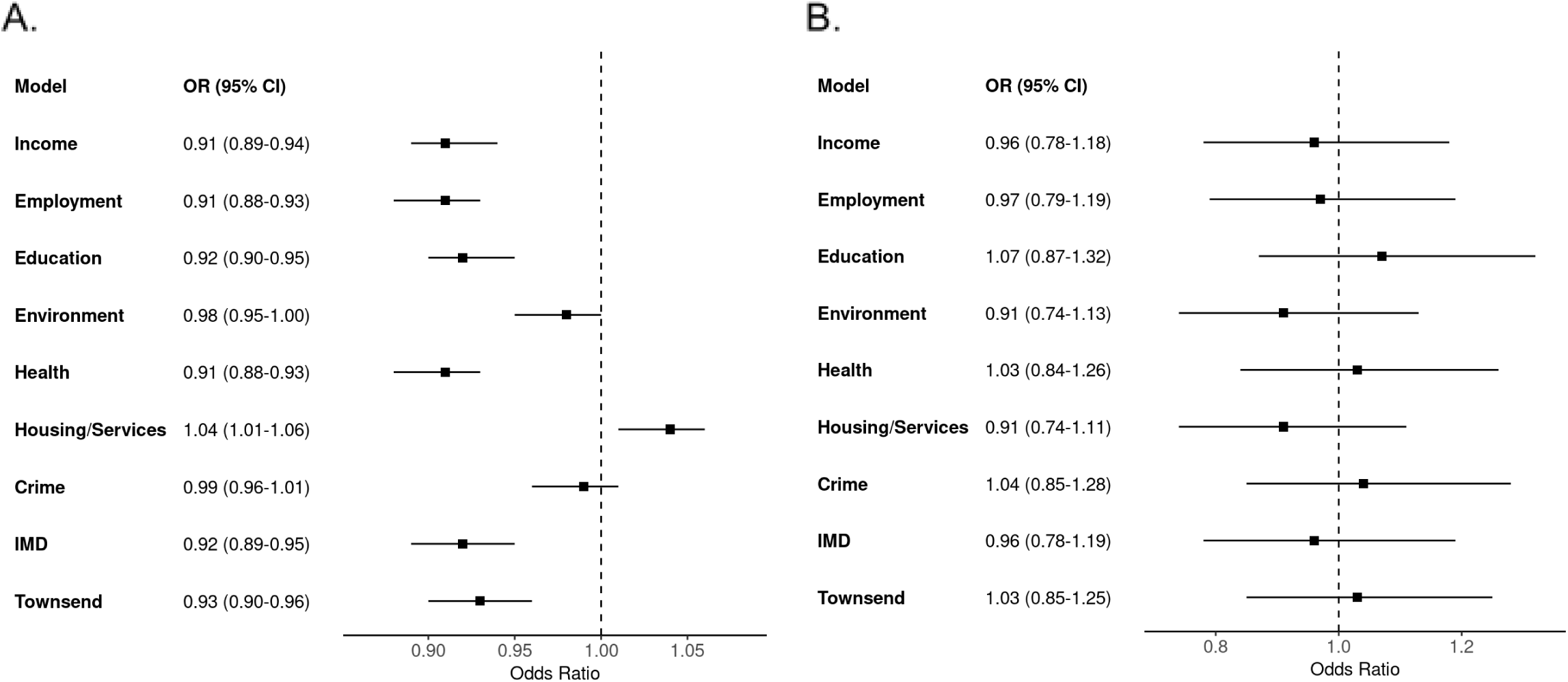
Odds ratios for prostate cancer in relation to nSES indices of. deprivation in (A) European group individuals and (B) African group individuals Odds ratios (ORs) and 95%confidence interval estimates between nSES deprivation indices and for the (A) European, and (B) African ancestries. All indices of deprivation, including index of multiple deprivation (IMD), were standardized per group prior to modeling. ORs<1 signify increased nSES deprivation and less neighborhood resources lower odds of prostate cancer

### PSA Screening and Healthcare Utilization Mediation

Differences in PSA screening and healthcare utilization may be mediating observed associations between nSES and PCa (22). Similar to the relationship between nSES deprivation and PCa, the Housing/Services index (OR=1.13; 95%CI =1.12-1.15; P<0.001) was positively associated with PSA screening in men of European ancestry. In this ancestry, the remaining eight indices were negatively associated with screening, with effects ranging from OR=0.93 (Housing and Education) to OR=0.97 (Environment). In the African ancestry models, the IMD, Income, Employment, Health, Education, and Townsend indices, were also negatively associated with ever having a PSA test. The remaining indices, including Housing/Services, were not associated with PSA screening (**Supp. Table 3**).

Inversely, all deprivation indices, except for Housing/Services (Beta=-0.33; 95%CI=-0.4--0.25; P<0.001), were positively associated with the number of clinical visits in men of European ancestry. The IMD (Beta=1.07; 95%CI=0.21-1.06; P=0.02), Health (Beta=1.06; 95%CI=0.10-2.02; P=0.03), Education (Beta=0.81; 95%CI=0.06-1.56; P=0.03), and Environment (Beta=1.00; 95%CI=0.16-1.84; P=0.02) indices were also positively associated with number of clinical visits in men of African ancestry (**Supp. Table 4**). When we calculated Pearson’s correlation between PSA screening, clinical visits, and the nSES indices, PSA screening was inversely correlated with all nSES indices while clinical visits were positively correlated with them. The exception in both cases was the Housing/Services index that had a correlation in the opposite direction (**Supp. Table 5**).

When included as covariates in the model between nSES deprivation and PCa, “Ever PSA Screened” was associated with PCa across models, while the number of clinical visits was not in any model in either ancestry. Additionally, the effect size of the nSES variables remained consistent after adjusting for the number of clinical visits and PSA screening status, including the statistically significant European ancestry associations: Education (OR=0.94; 95%CI=0.89-0.98; P=0.006), Employment (OR=0.91; 95%CI=0.86-0.96; P<0.001), Health (OR=0.91; 95%CI= 0.86-0.96; P<0.001), Income (OR=0.91; 95%CI=0.86-0.96; P<0.001), IMD (OR=0.92; 95%CI=0.87-0.96; P=0.001), and Townsend Index (OR=0.94; 95%CI=0.89-0.98; P=0.009) (**Supp. Table 6-8**).

Since PSA screening was associated with PCa when modeled with nSES, we undertook an additional analysis stratified by screening to reduce residual confounding remaining in the logistic regression model. In men of European ancestry, the association between nSES deprivation and prostate cancer remained consistent across screening status. For the never-screened individuals, the associations between nSES and PCa varied between OR=0.91 (IMD) to OR=0.94 (Townsend). We found no associations using the Crime and Environment index, and while not statistically significant, the Housing/Services index was the only variable with a positive association with PCa (OR=1.03; 95%CI=0.99-1.07; P=0.12)). In terms of the all screened group, IMD (OR=0.95; 95%CI=0.91-0.99; P=0.17), Income (OR=0.94; 95%CI=0.90-0.98; P=0.007), Employment (OR=0.94; 95%CI=0.90-0.98; P=0.003), Education (OR=0.95; 95%CI=0.92-1.00; P=0.32), and Townsend (OR=0.93; 95%CI=0.90-0.97; P=0.001) indices were also inversely associated with PCa. The nSES variable with the largest difference in effects was the Crime index (OR_never-screened_=0.97; 95%CI=0.93-1.01; P=0.10 & OR_screened_=1.01; 95%CI=0.97-1.06; P=0.43) (**Figure 2, Supp. Table 9**).

**Figure 2:**
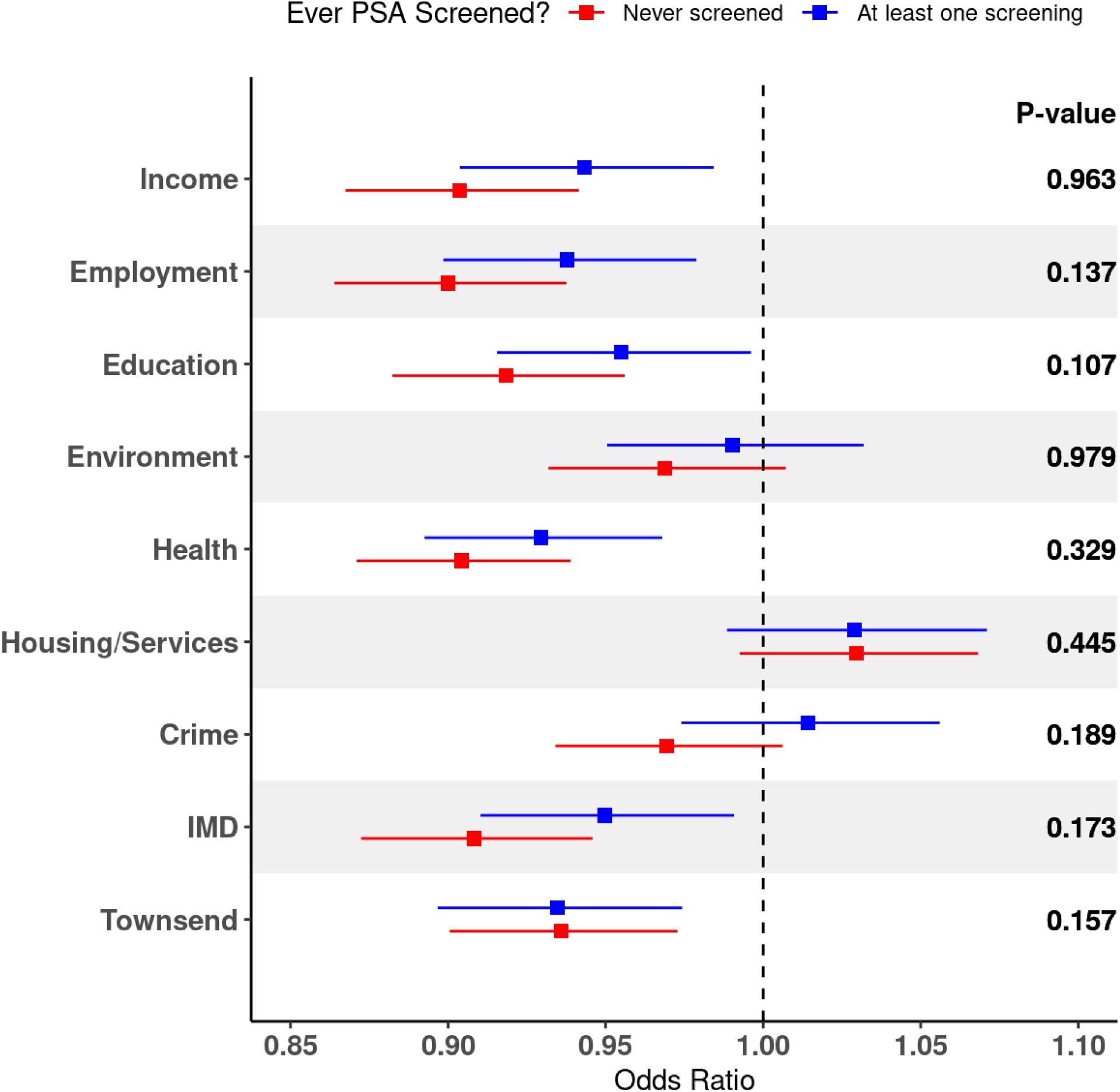
Odds ratios for prostate cancer in relation to nSES indices of deprivation stratified by PSA screening status. Odds ratios (ORs) and 95%confidence interval estimates between nSES deprivation indices and PCa for the European ancestry stratified by indicator for self-reported PSA screening at baseline (ever/never). All indices of deprivation, including index of multiple deprivation (IMD), were standardized prior to stratifying on screening status. P-values calculated as a difference of means z-test for each nSES variable stratified on PSA screening group. ORs<1 signify increased nSES deprivation and less neighborhood resources lower odds of prostate cancer

### PRS-by-nSES Effect Modification

When analyzing a potential GxE interaction between PRS_269_ and nSES deprivation, we found no effect modification across all indices for both the European (P>0.14) and African (P>0.56) ancestries (**Supp. Table 10**). To more thoroughly analyze if there were no GxE interactions, we compared the PRS_269_ association across nSES quartiles in men of European ancestry. When we did this, we found heterogeneity in the PRS_269_ associations across Townsend index quartiles (P=0.03) with associations ranging between OR=2.13 (Deprivation Quartile 1) to OR=1.93 (Deprivation Quartile 2) (**Table 3**). This analysis wasn’t performed in men of African ancestry due to the reduced power that would occur from stratifying an already small number of cases.

**Table 3:** Odds ratios for prostate cancer in relation to polygenic risk score (PRS) by neighborhood deprivation quartiles. Odds ratios (OR) and 95% confidence intervals for the association between continuous PRS_269_ values and PCa stratified by individuals in each neighborhood socioeconomic status (nSES) deprivation quartile for men of European ancestry. PRS_269_ values were standardized in the European ancestry sample prior to separating on deprivation quartile. Heterogeneity P-value calculated from a Cochran’s Q test comparing PRS_269_ associations within each stratified nSES deprivation index.

| **nSES Variables** | **Deprivation Q1**<br>**PRS Odds Ratio**<br>**(95% CI)** | **Deprivation Q2**<br>**PRS Odds Ratio**<br>**(95% CI)** | **Deprivation Q3**<br>**PRS Odds Ratio**<br>**(95% CI)** | **Deprivation Q4**<br>**PRS Odds Ratio**<br>**(95% CI)** | **Heterogeneity**<br>**p-value** |
| --- | --- | --- | --- | --- | --- |
| Index of Multiple Deprivation | 1.99<br>(1.89 - 2.1) | 2.12<br>(2 - 2.24) | 2.14<br>(2.02 - 2.26) | 2.03<br>(1.91 - 2.15) | 0.226 |
| Income Index of Deprivation | 2.03<br>(1.93 - 2.14) | 2.03<br>(1.92 - 2.15) | 2.15<br>(2.04 - 2.28) | 2.06<br>(1.94 - 2.19) | 0.418 |
| Employment Index of Deprivation | 2.04<br>(1.94 - 2.15) | 2.05<br>(1.94 - 2.16) | 2.17<br>(2.06 - 2.3) | 2.01<br>(1.89 - 2.13) | 0.231 |
| Health Index of Deprivation | 2.03<br>(1.92 - 2.14) | 2.14<br>(2.03 - 2.26) | 2.12<br>(2 - 2.24) | 1.97<br>(1.86 - 2.1) | 0.159 |
| Education Index of Deprivation | 2.1<br>(1.99 - 2.21) | 2.03<br>(1.92 - 2.15) | 2.06<br>(1.95 - 2.18) | 2.08<br>(1.95 - 2.2) | 0.857 |
| Housing/Services Index of Deprivation | 2.03<br>(1.92 - 2.15) | 2.06<br>(1.94 - 2.17) | 2.02<br>(1.91 - 2.14) | 2.16<br>(2.04 - 2.29) | 0.324 |
| Crime Index of Deprivation | 2.01<br>(1.91 - 2.13) | 2.02<br>(1.91 - 2.13) | 2.19<br>(2.07 - 2.32) | 2.06<br>(1.94 - 2.18) | 0.115 |
| Environment Index of Deprivation | 1.98<br>(1.88 - 2.09) | 2.11<br>(2 - 2.23) | 2.09<br>(1.98 - 2.22) | 2.1<br>(1.98 - 2.22) | 0.37 |
| Townsend Index of Deprivation | 2.13<br>(2.02 - 2.25) | 1.93<br>(1.83 - 2.03) | 2.1<br>(1.99 - 2.23) | 2.12<br>(2 - 2.25) | 0.029 |

## DISCUSSION

We investigated potential main effects and interactions between a well-established risk factor, germline genetics, and a less well-studied exposure, nSES as measured through deprivation, and PCa.

Previous studies analyzing nSES and PCa reported conflicting results with some finding a positive association (7,9,24) and others finding a negative association (8,10,25). These variations are likely due to differences in study population, methodology, country of study, healthcare systems, and utilization of different measures of nSES deprivation and covariates. And while the association between nSES and PCa incidence is unclear, studies consistently found nSES deprivation positively associated with both PCa stage and mortality (4,6,9). Our findings align with the subset of studies, performed in the United States, Taiwan, and Scotland that find increasing neighborhood resources (or decreasing deprivation) increases PCa risk, specifically in the domains of education, employment, health, and income (9,24,25). These studies suggest that higher nSES groups are more likely to be PSA screened or utilize the healthcare system (26). While PSA screening was positively associated with PCa, healthcare utilization was not. Moreover, nSES was associated with PCa after adjusting for self-reported screening at baseline. One possible explanation for the nSES association is that while PSA screening may lead to overdiagnosis, there may be environmental exposures more common in resourced neighborhoods that increase PCa incidence (2). More research investigating environmental factors associated with nSES and PCa risk is needed to further investigate this.

The only nSES deprivation domain positively associated with PCa was the Housing/Services index, implying that men in neighborhoods with less access to housing and services were more likely to be diagnosed with PCa. The mechanism underlying this association, and its opposite effect from other domains, is unknown. It is likely connected to our finding that the Housing/Services index is uncorrelated with the other indices, except for the Environment and Townsend indices that include housing deprivation in their calculation. When we remove housing-relevant indicators, the Housing/Services index is calculated using the average distance to key services, such as intensive medical care, supermarkets, schooling, and post offices. The association between Housing/Services and PCa is likely driven by services deprivation, particularly factors associated with distance to medical care and schooling. The associations between nSES deprivation and PCa were also only seen in the European ancestry group. Due to the smaller number of African ancestry men, it is difficult to ascertain if there is no effect in these groups or if we were underpowered to identify these associations.

We also reaffirm the relationship between PCa PRS and PCa risk in a sample of European ancestry men in UKB using a 269 variant PRS model. As previously shown, the PRS association with PCa increased over PRS deciles (14,27,28). These associations and trends were diminished in the African ancestry. Attenuated PRS associations for non-European ancestries, including African ancestries, have been consistently replicated across PCa PRS models (3,19,29,30). Current explanations include differences in linkage disequilibrium patterns, allelic effect size, and causal allele frequency whose effects are exacerbated by smaller African ancestry sample sizes in the GWAS that generate PRS models (19). The lack of PRS associations in men of African ancestry is likely attributed to the smaller sample sizes of this population in the UKB.

Gene-environment (GxE) interactions are thought to be driven by biological processes or spurious associations, and are of increasing interest for improving genetic discovery and explaining missing heritability and population heterogeneity (31). When we investigated the potential role of gene-nSES interactions to explain PCa heritability and heterogeneity in previous studies, we identified an interaction between PRS_269_ and the Townsend index. The lack of an interaction for the other index measuring overall deprivation, the Index of Multiple Deprivation, may signify that the interaction with Townsend index is a spurious association. While GxE interactions are challenging to detect due to limited statistical power, the absence of interactions for the other indices may reflect that nSES and PRS act independently. It could also reflect differences in factors affecting PCa development versus those affecting PCa detection. There may be environmental factors that modify genetic susceptibility to PCa, but more work must be done to identify variables that are risk factors themselves for PCa. Future work should also incorporate novel frameworks and techniques to facilitate the discovery of these interactions and explain the remaining variation in PCa risk (31,32).

Several key limitations prevent us from interpreting our results more broadly. First, the limited number of African ancestry individuals in UKB, combined with the known decrease in PRS performance for ancestries underrepresented in the training data (30,33), limited our ability to detect effect modification and identify PRS association trends in men of African ancestry. Future studies require larger cohorts that are more diverse in ancestry, personal identity, and socioeconomic background (34). Second, we lacked in-depth cancer stage and screening information to determine the magnitude of our PCa associations and PSA screening effect. For example, our PSA screening variable was a binary self-reported measurement taken at assessment. To avoid measurement error, reporting bias, and other confounding, a more-precise, repeated, and validated variable was required. Lastly, nSES indices are broad measures of deprivation assigned at assessment without considering how long they lived in these neighborhoods. This could lead to exposure misclassification for nSES in the etiologically relevant time frame prior to PCa diagnosis. In addition, individual SES, which varies within geographic regions, can add unmeasured confounding to the nSES relationship studied in this research (24,35). Given available data, it was not possible to adjust for this confounding across indices measuring domain-specific deprivation and overall deprivation in a principled manner. While previous research has adjusted for individual SES and identified associations with nSES (8,9), future work should account for an individual’s SES in specific socioeconomic domains and length of exposure in a specific neighborhood.

In conclusion, we identified an inverse relationship between several domains of neighborhood deprivation and PCa while accounting for potential mediators, such as PSA screening and healthcare utilization. As the first study to examine domains of nSES in PCa, this research provides an opportunity to investigate non-genetic variation in PCa risk. Additionally, we reaffirmed the association between germline genetics and PCa while attempting to identify novel gene-by-nSES interactions. While such interactions were not identified, more research focused on other exposures, specifically those that modify disease development and not only disease detection, is necessary to improve our knowledge of PCa etiology.

## Data Availability Statement

The research was conducted with approved access to UK Biobank data under application number 14105 (PI: Witte). UK Biobank data are publicly available by request from https://www.ukbiobank.ac.uk. Informed consent was obtained from all study participants. UK Biobank received ethics approval from the Research Ethics Committee (REC reference: 11/NW/0382). PRS_269_ (PRS ID: PGS000662) and PRS_451_ (PRS ID: PGS003765) are publicly available on the PGS catalog from https://www.pgscatalog.org/.

## Data Availability

The research was conducted with approved access to UK Biobank data under application number 14105 (PI: Witte). UK Biobank data are
publicly available by request from https://www.ukbiobank.ac.uk. Informed consent was
obtained from all study participants. UK Biobank received ethics approval from the
Research Ethics Committee (REC reference: 11/NW/0382). PRS269 (PRS ID:
PGS000662) and PRS451 (PRS ID: PGS003765) are publicly available on the PGS
catalog from https://www.pgscatalog.org/.

## Acknowledgements

The funder had no role in the design of the study; the collection, analysis, or interpretation of the data; or the writing of the manuscript and decision to submit it for publication.

